# Optimising the construction of outcome measures for impact evaluations of intimate partner violence (IPV) prevention interventions

**DOI:** 10.1101/2023.02.07.23285510

**Authors:** Sangeeta Chatterji, Christopher Boyer, Vandana Sharma, Tanya Abramsky, Ruti Levtov, Kate Doyle, Sheila Harvey, Lori Heise

## Abstract

Most impact evaluations of IPV prevention interventions use binary measures of “any” versus “no” physical and/or sexual IPV as their primary outcome measure, missing opportunities to capture nuance. In this study, we reanalysed secondary data from six randomised controlled trials conducted in low and middle-income countries- Bandebereho (Rwanda), Becoming One (Uganda), Indashyikirwa (Rwanda), MAISHA CRT01, MAISHA CRT02 (Tanzania), Stepping Stones Creating Futures (South Africa), and Unite for a Better Life (Ethiopia), to assess how different conceptualisations and coding of IPV variables can influence interpretations of the impact of an intervention. We compared standard outcome measures to new measures that reflect the severity and intensity of violence and whether interventions prevent new cases of IPV or reduce or stop ongoing violence. Results indicate that traditional binary indicators masked some of the more subtle intervention effects, and the use of the new indicators allowed for a better understanding of the impacts of the interventions. Conclusions on whether a program is perceived “to work” are highly influenced by the IPV outcomes investigators choose to report and how they are measured and coded. Lack of attention to outcome choice and measurement could lead to prematurely abandoning strategies useful for violence reduction or missing essential insights into how programs may or may not affect IPV. While these results must be interpreted cautiously, given differences in intervention types, the underlying prevalence of violence, sociodemographic factors, sample sizes and other contextual differences across the trial sites, they can help us move toward a new approach to reporting multiple outcomes that allow us to unpack the ‘impact’ of an intervention by assessing intervention effect by the severity of violence and type of prevention, whether primary and secondary.

## Introduction

Over the last decade, more than 95 randomised controlled trials and quasi-experimental evaluations have been conducted on intimate partner violence (IPV) prevention interventions (Dickens et al., 2019; Kerr-Wilson et al., 2019). This knowledge base provides us with a unique opportunity to review methodological and measurement issues of particular relevance to the field of violence prevention. Early scholarship on violence and measurement focused on capturing accurate IPV prevalence data, including optimising the construct and content validity of measures (Follingstad & Rogers, 2013; Hardesty et al., 2015; Waltermaurer, 2005), assessing participation bias (McNutt & Lee, 2000; Waltermaurer et al., 2003), maximising the precision of estimates and quality of data (Lehrner & Allen, 2014; Ruiz-Pérez et al., 2007), and exploring inconsistency and gender differences in disclosure (Chan, 2011; Hamby, 2016; Rowlands et al., 2020; Straus, 2017) among other issues (Bender, 2016; Follingstad & Rogers, 2013; Hamby, 2005; Ruiz-Pérez et al., 2007). More recently, researchers have assessed the equivalence of IPV scales across countries (Yount et al., 2022) and developed suggested thresholds for coding the severity of emotional/psychological aggression (Heise et al., 2019).

Much of this research has been conducted within the field of psychology. By contrast, methodological research of special relevance to evaluating the *impact* of prevention interventions has lagged behind. Researchers have begun to address this gap by developing ways to assess whether an intervention prevents new cases of violence and/or reduces the frequency of violence already underway at baseline (Chatterji et al., 2020). Likewise, other investigators have assessed the measurement invariance of various IPV outcome measures between baseline and endline and across the arms of various IPV prevention trials (Clark et al., 2022). In this paper, we build on this growing body of methodological work by assessing how different ways of defining and coding IPV in prevention trials influence our interpretation of how (and whether) different interventions may work to reduce IPV.

### Background

In evaluation research, conclusions about the success of an intervention depend on an assessment of one or more primary outcomes. Consequently, how these variables are coded affects the inferences we draw from our data. Increasing measurement precision allows us to develop nuanced constructs to answer more complex conceptual questions about violence (Grych & Hamby, 2014). Most impact evaluations of IPV prevention interventions use binary measures of “any” versus “no” physical and/or sexual IPV as their primary outcome measure, missing opportunities to capture nuance. More recently, a review found that some trialists have begun to report on a broader range of outcomes, offering separate estimates of how an intervention impacts physical, sexual, and emotional IPV (Keith et al., 2022). Reporting on multiple types of IPV allows for a better understanding of the impact of an intervention, as different types of IPV are distinct from one another, and interventions may impact one or more forms of violence.

It is likewise essential to assess intervention impact by the severity of violence. Studies that categorise acts of physical IPV by severity, have found that severe acts are associated with more negative health outcomes (Lacey & Mouzon, 2016; Signorelli et al., 2014; Smith et al., 2010) and a higher risk for future perpetration of more severe violence (Cunha & Goncalves, 2018). Similarly, there is evidence of a dose-response relationship between the intensity of emotional IPV and adverse health outcomes (Heise et al., 2019). Although there is enough evidence to show that severe violence is associated with more negative health outcomes, there is no consensus on the types of acts considered severe, the threshold of severity, or the best way to measure severity. Studies that have examined IPV severity used the Revised Conflict Tactics Scale (CTS2) (Korman et al., 2008; Smith et al., 2010; Straus et al., 1996), a single-item measure (Lacey et al., 2020; Lacey & Mouzon, 2016), a continuous measure to assess severity linearly (Ferrari et al., 2014) or latent class analysis to develop thresholds of severity (Heise et al., 2019). The Conflict Tactics Scale (CTS), the most widely used instrument to measure IPV and severe IPV (Bender, 2016), was one of the first measures that categorised acts of physical IPV into two classes, minor and severe physical IPV (Straus, 1979). Two decades later, the revised CTS2 included more types of IPV and differentiated between minor and severe acts of physical assault, sexual coercion, and psychological aggression (Straus et al., 1996). These severity classifications are driven by types of injury and other health consequences of violence.

In evaluation research, IPV interventions can differ in their impacts on primary versus secondary prevention. Primary prevention works by preventing violence before it occurs; secondary prevention works by reducing or stopping ongoing abuse (Ellsberg et al., 2015). For example, in primary trial analyses, the *Indashyikirwa* intervention in Rwanda impacted physical and sexual IPV among all women (Dunkle et al., 2020). Subsequent analysis demonstrated that the intervention worked by reducing and/or stopping ongoing physical and sexual IPV among women reporting violence at baseline. The intervention was ineffective at preventing the onset of IPV or primary prevention among women who did not report ongoing IPV at baseline (Chatterji et al., 2020). In another study, *SASA!,* a community-based intervention in Uganda, was slightly more effective at reducing ongoing sexual and physical IPV, than at preventing the onset of these types of IPV (Abramsky et al., 2016). These differences in program impact are only evident when we conduct further analyses to assess the differential impact of an intervention on primary versus secondary prevention. Such distinctions can help trialists and practitioners engage the most appropriate populations for a particular intervention.

In this paper, we build on this work by reanalysing trial data to assess how different conceptualisations and coding of IPV variables can influence interpretations of the impact of an intervention. We hope this exercise will initiate a discussion on the broader violence prevention community on the trade-offs of reporting multiple IPV outcomes and their conceptualisation to advance our understanding of IPV measurement and identify new directions for future research. We use secondary data from six randomised controlled trials conducted in low and middle-income countries--*Bandebereho, Becoming One, Indashyikirwa, MAISHA CRT01, MAISHA CRT02, Stepping Stones Creating Futures,* and *Unite for a Better Life,* to compare different ways to code IPV outcomes and assess any potential differences in measured effectiveness of the interventions based on IPV severity or type of IPV prevention.

## Methods

### Description of Studies

Appendix Table 3 provides an overview of the six different trials included in this paper. The *Bandebereho* trial in Rwanda was a two-arm multi-site randomised controlled trial. The intervention uses the transition to parenthood as an entry point to work with men and their partners to transform harmful masculine attitudes and support more equitable and non-violent couple and family relationships. A 15-session curriculum covers topics such as gender and power, fatherhood; couple communication and decision-making; IPV; child development; and men’s engagement in prenatal and infant care. Men participated in all sessions, and women in up to 8 sessions. At 21 months, 94% of men (1123) and 97% of women (1162) were retained (Doyle et al., 2018). Data from female participants is used for the secondary analyses presented in this study.

*Becoming One* was evaluated using an individually randomised controlled trial in Uganda. In this intervention, faith leaders take groups of couples through a 12-week curriculum designed to strengthen their relationship and prevent or reduce IPV. At baseline, 1680 couples were assigned to intervention and control groups, and endline retention at 12 months was 100%. The intervention improves couples’ relationships by leveraging the church’s authority to shift perceptions of norms surrounding proper behaviour in relationships. It includes sessions on communication, conflict-resolution skills, negotiating consent and desire, sharing financial responsibilities and re-interpretation of biblical passages (Boyer et al., 2022).

The *Indashyikirwa* (Agents for change) trial in Rwanda was a community-randomised controlled trial. The intervention includes four interlocking components, a 21-session couples’ curriculum, activist training and community activism, opinion leader training, and women’s safe spaces. The couples’ curriculum, an intensive gender transformative and relationship-strengthening intervention, addressed positive and negative types of power, critical triggers of IPV (i.e. jealousy, alcohol abuse, economic stress), and skills building around communication and conflict resolution. At 24 months, 97% of women (1617) and 93% of men (1536) were retained (Dunkle et al., 2020).

*MAISHA CRT01* evaluated the MAISHA curriculum (Wanawake Na Maisha) using a cluster-randomised controlled trial in Tanzania. Women participating in a microfinance loan scheme were invited to participate in a social empowerment program where they developed skills to minimise and prevent IPV and defend themselves against it and its negative consequences. Topics included knowledge and awareness of traditional gender norms and IPV, communication and conflict resolution skills, peer support and social capital. At 24 months, 89% (485) of the intervention and 86% (434) of the control group women provided data for the impact evaluation (Kapiga et al., 2019). The *MAISHA CRT02* trial evaluated the impact of the same intervention on the IPV experiences of women residing in the same neighbourhoods which were not part of any microfinance groups. At the 24-month follow-up, 88% (551) of intervention and 90% (575) of the control group women provided data (Harvey et al., 2021).

The *Stepping Stones and Creating Futures (SS-CF)* trial in South Africa was a two-arm cluster randomised controlled trial with a wait-list control condition. *SS-CF* is a behavioural intervention to reduce IPV by transforming gender attitudes and relationships and strengthening livelihoods. Women and men were included in the study in separate groups, and these participants were typically not in romantic relationships with one another. At 24 months, endline retention was 74.9% (505) for men and 80.6% (545) for women (Gibbs et al., 2020).

*Unite for a Better Life* was evaluated using a cluster-randomized controlled trial in Ethiopia. UBL is a participatory gender-transformative intervention delivered to groups of women, men, or couples during the Ethiopian coffee ceremony, a cultural forum for discussion and reflection. The intervention addressed the root causes of gender-based inequalities by examining and challenging traditional gender norms and power imbalances during 14 facilitator-led skill-building sessions. Topics included gender norms, sexuality, communication and conflict resolution, HIV/AIDS, and IPV. At 24 months post-intervention, 88% of trial participants surveyed at baseline (5248) and 87% of their spouses (5131) provided follow-up data (Sharma et al., 2020). This paper used men’s and women’s data from the men’s UBL group and the control group for secondary analysis.

### Measures

To explore how the choice of coding affects the measured impact of each intervention, **w**e constructed a range of new outcome measures for the reanalysis of data from the trials above. All trials used a version of the WHO instrument for assessing IPV (García-Moreno, Jansen, Ellsberg, Heise, & Watts, 2005). The original WHO study included measures of physical IPV (5 acts), sexual IPV (3 acts), and emotional IPV (4 acts). All scales used behaviourally specific questions to inquire about women’s victimisation and men’s perpetration of IPV over the past 12 months (e.g., in the past 12 months, how many times has a current husband or boyfriend ever slapped you or thrown something at you which could hurt you?). Responses typically were: ‘0=never’, ‘1=once’, ‘2=a few times’, or ‘3=many times.’ Appendix table 4 presents the items used in different trials.

### Physical IPV

Investigators traditionally coded physical IPV as a binary variable, with a “case” of physical IPV defined as anyone who has experienced or perpetrated one or more of the physical acts of violence included in the WHO or DHS instruments. We compared this measure to two new measures that distinguished between moderate and severe physical violence. The physical IPV items were divided into moderate and severe acts of physical IPV, as defined and validated in the CTS2 (Straus et al., 1996). A participant was coded as having experienced/perpetrated moderate only physical IPV if they experienced at least one of the two acts of moderate physical IPV (slapping or throwing something that could hurt the participant; pushing or shoving the participant, or pulling the participant’s hair) at any frequency, and did not experience/perpetrate any acts of severe physical IPV. Severe physical IPV included participants who had experienced/perpetrated any of the four acts of severe physical IPV (hit with a fist or something else that could hurt; kicked, dragged, beat up; choked or burnt; threatened to use a weapon or used weapon) at any frequency.

### Severe physical and/or sexual IPV

We compared two measures of ‘severe’ physical and/or sexual IPV. In the first measure of severe physical and/or sexual IPV, participants were coded as a ‘case of severe IPV’ if they reported any of the four items of severe physical IPV (hit with a fist or something else that could hurt; kicked, dragged, beat up; choked or burnt; threatened to use a weapon or used weapon) or any item measuring sexual IPV at any frequency. The second measure of severe physical and/or sexual IPV uses the approach of the What Works to prevent Violence Program. The What Works to Prevent Violence Against Women and Girls program, was an 8-year research collaboration funded by the UK government between 2012-20. Fifteen interventions were developed and evaluated for this program in LMICs. *Indashyikirwa* and *Stepping Stones Creating Futures* were part of the What Works program. This measure includes the experience/perpetration of any act of physical IPV or sexual IPV more than once (a few or many times in frequency) or the experience/perpetration of two or more different types of physical or sexual IPV at any frequency (Dunkle et al., 2020).

### Emotional IPV

Typically, emotional IPV variables measure the experience of any act of emotional IPV at any frequency. We compared this measure to two new approaches to estimating the intensity of emotional IPV. These measures are based on preliminary results of measurement equivalence and latent class analysis from another study on the measurement of emotional abuse for global reporting on Sustainable Development Goals (SDGs) (Clark et al. *under review*).

We first created a variable that measures three categories of emotional IPV based on the act type and frequency. For sites that included three items to measure emotional IPV:

- High-intensity emotional IPV includes individuals who report experiencing/perpetrating both insults and humiliation “often/many times” or experiencing/perpetrating threats “often/many times”.
- Moderate-intensity emotional IPV includes individuals who report experiencing/perpetrating insults and humiliation “sometimes/a few times” or threats “sometimes/a few times”.
- Low or no emotional IPV includes all other experiences. For sites that had four items for emotional IPV:
- High-intensity emotional IPV includes individuals who report experiencing/perpetrating at least two of the acts of insults, humiliation/belittling, and scaring “often/many times” or experiencing/perpetrating threats “often/many times” alone.
- Moderate-intensity emotional IPV includes individuals who report experiencing/perpetrating at least two acts of insults, humiliating/belittling, and scaring “sometimes” or experiencing/perpetrating threats “sometimes” alone.
- Low or no emotional IPV includes all other experiences.

This 3-level variable was then recoded to create two binary variables: 1) moderate and/or high-intensity emotional IPV vs low or no emotional IPV; and 2) high-intensity emotional IPV only vs low or no emotional IPV.

### Primary vs secondary prevention

To assess differences in treatment outcomes by baseline reporting of IPV, we used three binary variables: cessation, reduction, and prevention, tested in a prior study (Chatterji et al., 2020).

Among individuals who *<u>reported</u>* past-year experience/perpetration of IPV at baseline, *reduction* assesses whether IPV reduced between baseline and endline (1=IPV reduced at endline, 0=IPV stays the same/increased between baseline and endline). Among individuals who reported past-year experience/perpetration of IPV at baseline, *cessation* measures whether IPV stopped completely between baseline and endline (1=IPV stopped at endline, 0= IPV stays the same/increased/reduced but did not stop entirely between baseline and endline).

Among individuals who did *not* report experiencing/perpetrating any given type of violence at baseline*, prevention* evaluates whether the intervention stopped new cases of IPV from occurring during follow-up (1=participants continued reporting no IPV experience/perpetration at endline, 0=participant reported experiencing/perpetrating IPV at endline).

### Analysis

All trials used an intention-to-treat approach. We conducted our secondary analysis of trial findings using new outcome measures using the same modelling strategy as the primary authors used for the original trial. These secondary analyses were not pre-specified for any sites and should be considered exploratory.

In the *Bandebereho* trial, outcomes were analysed using generalised estimating equations accounting for the clustered nature of the data (Doyle et al., 2018). *Becoming One* used least-squares regression that conditions an indicator for the treatment assignment, fixed effects for the pair blocks, and covariates selected through a cross-validated lasso regression to assess the effectiveness of the intervention (Boyer et al., 2022). In the *Indashyikirwa* trial, outcomes were analysed using generalised linear mixed-effects models with a logit link function to compare the effect of the intervention between the two study arms for all binary variables (Dunkle et al., 2020). *MAISHA CRT01’s* impact was assessed using logistic regression models with a random intercept for the microfinance group to account for the clustered data (Kapiga et al., 2019). *MAISHA CRT02* also employed the same models with random intercepts for neighbourhood clusters (Harvey et al., 2021). Outcomes for the *SS-CF* trial were analysed using generalised estimating equation models accounting for the clustered nature of the data (Gibbs et al., 2020). *UBL’s* impact was also assessed using logistic regression models fitted with generalised estimating equations with strata-fixed effects for district and standard errors clustered at the village-level (Sharma et al., 2020). Except for *Bandebereho* and *UBL,* where baseline data were unavailable, study samples were stratified by baseline reporting of IPV experience to assess the differential impact on primary and secondary prevention. This analysis was conducted for *Indashyikirwa* and *SS-CF* in a prior study (Chatterji et al., 2020) and not reported here. We report 95% confidence intervals and *p-*values for all outcomes. Analysis was conducted using Stata version 16.

### Ethical approval

The *Bandebereho* study received ethical approval from the Rwanda National Health Research Committee, the Rwanda National Ethics Committee, and the Rwandan National Institute of Statistics (Doyle et al., 2018). Ethical approval for the *Becoming One* study was obtained from Innovations for Poverty Action, the Mildmay Uganda Research and Ethics Committee, and the Ugandan National Council for Science and Technology (Boyer et al., 2022). Ethical approval for the Indashyikirwa study was obtained from the Rwandan National Ethics Committee, the National Institute of Statistics Rwanda and the South Africa Medical Research Council (Dunkle et al., 2020). *MAISHA CRT01* and *MAISHA CRT02* obtained ethical approval from the Tanzanian National Health Research Ethics Committee of the National Institute for Medical Research and the London School of Hygiene and Tropical Medicine ethics committee (Harvey et al., 2021). Approval to undertake the *SS-CF* trial was granted by the ethics committees of the University of KwaZulu-Natal, Durban, South Africa and the South African Medical Research Council Ethics Committee (Gibbs et al., 2020). Approval to conduct the *UBL* trial was sought from the Committee on the Use of Humans as Experimental Subjects at the Massachusetts Institute of Technology and the IRB board at the Addis Ababa University College of Health Sciences (Sharma et al., 2020). Written consent was obtained from participants at all but one site; illiterate participants could have the form read to them by study personnel or a trusted person of their choosing. *UBL* obtained oral consent from all participants (Sharma et al., 2020).

## Results

### Descriptive data

#### Physical IPV

Table 1 presents descriptive and multivariate results. Across all sites, most women who disclosed any physical IPV reported experiencing severe acts of violence, far fewer experienced only moderate acts of violence. For example, in *UBL*, 13.1% of women reported experiencing severe acts of violence, compared to 20% experiencing any physical acts. A significantly smaller proportion, 6.7% of women, experienced moderate only physical violence. Similarly, in *Becoming One*, 14.9% of women experienced severe physical violence compared to 8.9% reporting moderate only physical violence and 23.8% reporting any physical violence.

**Table 1:**
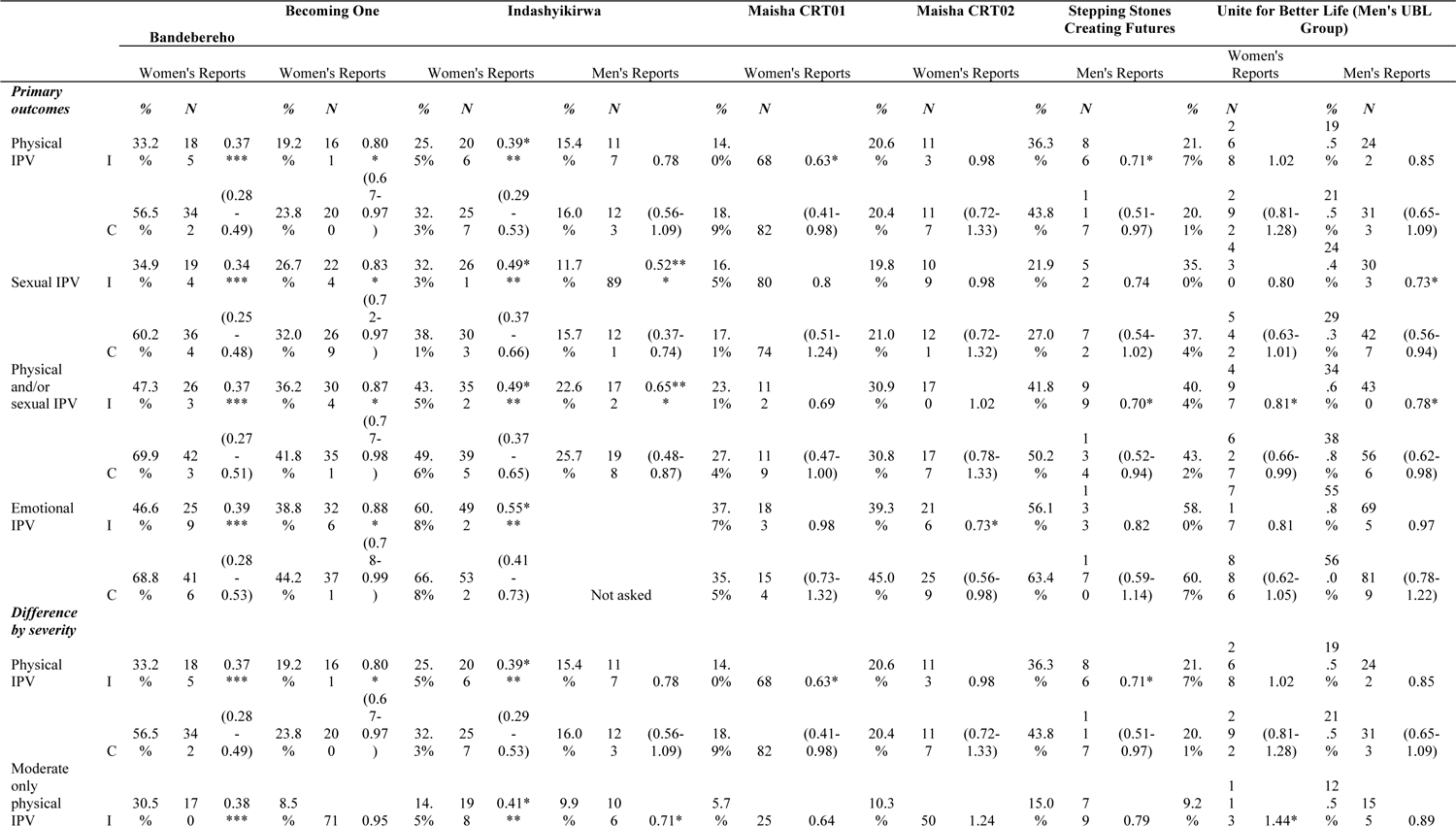
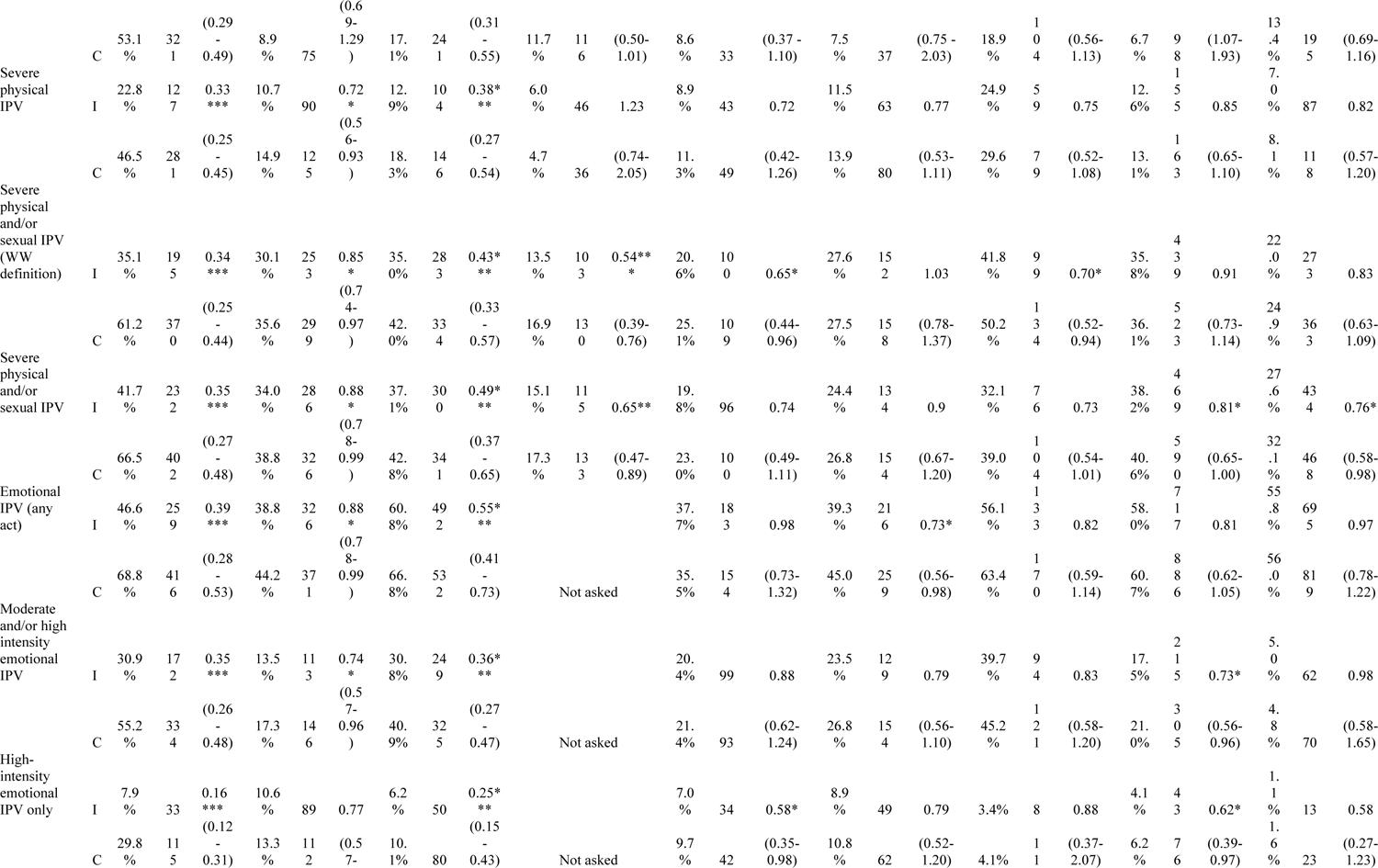
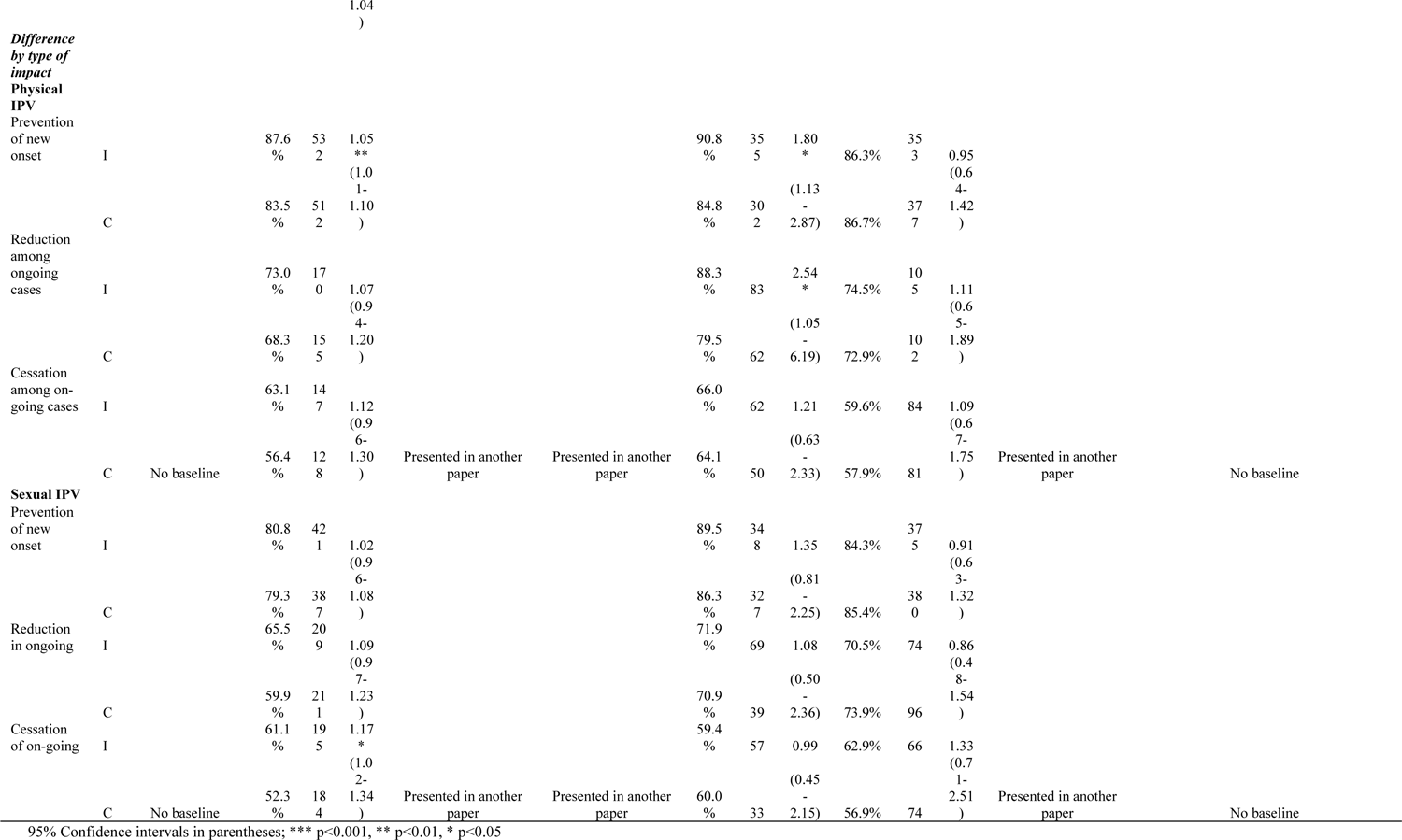
Examining differences in intervention impact using different measurement & coding practices

**Table 2:** Examining differences in intervention impact using different measurement & coding practices-results presented visually

| Variable | Bandebereho | Becoming One | Indashyikirwa |  | Maisha 1 | Maisha 2 | SS Creating Futures | Unite for Better Life (Men's Only Group) |  |
| --- | --- | --- | --- | --- | --- | --- | --- | --- | --- |
|  | Women's Reports | Women's Reports | Women's Reports | Men's Reports | Women's Reports | Women's Reports | Men's Reports | Women's Reports | Men's Reports |
| **Primary outcomes** |  |  |  |  |  |  |  |  |  |
| Physical IPV | Sig | Sig | Sig | Not sig | Sig | Not sig | Sig | Not sig | Not sig |
| Sexual IPV | Sig | Sig | Sig | Sig | Not sig | Not sig | Not sig | Not sig | Sig |
| Physical and/or sexual IPV | Sig | Sig | Sig | Sig | Not sig | Not sig | Sig | Sig | Sig |
| Emotional IPV | Sig | Sig | Sig | Not sig | Not sig | Sig | Not sig | Not sig | Not sig |
| **Difference by severity** |  |  |  |  |  |  |  |  |  |
| Physical IPV | Sig | Sig | Sig | Not sig | Sig | Not sig | Sig | Not sig | Not sig |
| Moderate only physical IPV | Sig | Not sig | Sig | Sig | Not sig | Not sig | Not sig | Sig | Not sig |
| Severe physical IPV | Sig | Sig | Sig | Not sig | Not sig | Not sig | Not sig | Not sig | Not sig |
| Severe physical and/or sexual IPV (WW definition) | Sig | Sig | Sig | Sig | Sig | Not sig | Sig | Not sig | Not sig |
| Severe physical and/or sexual IPV | Sig | Sig | Sig | Sig | Not sig | Not sig | Not sig | Sig | Sig |
| Emotional IPV (any act) | Sig | Sig | Sig | Not asked | Not sig | Sig | Not sig | Not sig | Not Sig |
| Moderate and/or high-intensity emotional IPV | Sig | Sig | Sig | Not asked | Not sig | Not sig | Not sig | Sig | Not Sig |
| High intensity emotional IPV only | Sig | Not sig | Sig | Not asked | Sig | Not sig | Not sig | Sig | Not Sig |
| **Difference by type of impact** |  |  |  |  |  |  |  |  |  |
| **Physical IPV** |  |  |  |  |  |  |  |  |  |
| Prevention of new onset | No baseline | Sig | Results presented in another paper |  | Sig | Not sig | Results presented in another paper | No baseline |  |
| Reduction among ongoing cases | No baseline | Not sig |  |  | Sig | Not sig |  | No baseline |  |
| Cessation among on-going cases | No baseline | Not sig |  |  | Not sig | Not sig |  | No baseline |  |
| **Sexual IPV** |  |  |  |  |  |  |  |  |  |
| Prevention of new onset | No baseline | Not sig |  |  | Not sig | Not sig |  | No baseline |  |
| Reduction in ongoing | No baseline | Not sig |  |  | Not sig | Not sig |  | No baseline |  |
| Cessation of on-going | No baseline | Sig |  |  | Not sig | Not sig |  | No baseline |  |

The results are different for men, with a higher proportion of men reporting perpetrating moderate only physical IPV as compared to severe physical IPV. In *UBL*, 13.4% of men in the control group reported perpetrating moderate only physical violence, 8.1% severe physical violence and 21.5% any physical violence. In *Indashykirwa*, 11.7% of men reported perpetrating moderate only physical violence, 4.7% severe physical violence, and 16% any physical violence.

#### Severe physical and/or sexual IPV

The prevalence of severe physical and/or sexual violence differed by how the outcome was coded. In *MAISHA CRT01*, 23% of women reported experiencing severe physical and/or sexual violence as per the first measure, and 25% severe physical and/or sexual violence per the What Works measure.

The results were similar for male participants; in *UBL*, 32% of men in the control group reported perpetrating any severe physical and/or sexual violence, and 24.9% using the What Works measure of severe physical and/or sexual violence.

#### Emotional IPV

Most respondents experienced moderate and/or high-intensity emotional IPV as compared to high-intensity emotional IPV only. In the *Bandebereho* trial, 55.2% reported moderate and/or high-intensity emotional violence, and 29.8% reported high-intensity only emotional violence. When using the traditional outcome measure, 68.8% reported experiencing any emotional violence.

Among male participants in the control group in the *UBL* intervention study, 4.8% reported perpetrating moderate and/or high-intensity emotional violence, and 1.6% high-intensity emotional IPV only. Using the traditional outcome measure, 56% reported perpetrating any emotional violence.

### Differences in intervention impact by levels of severity

#### Physical IPV

Interventions differed in their impact on physical IPV when compared by category of severity. In the *Becoming One* trial, the intervention impacted women’s experiences of physical IPV (aRR:0.80, C.I.: 0.67-0.97) when analysing the traditional indicator. However, the intervention did not impact moderate only physical IPV but did have a significant impact on severe physical IPV (aRR:0.72, C.I.: 0.56-93). Conversely, the men’s *UBL intervention* had no effect on women’s reported experience of any physical IPV. However, the current study findings suggest the intervention may have increased women’s reports of moderate only physical IPV (aOR:1.44, C.I.: 1.07-1.93) but had no effect on severe only physical IPV. *Bandebereho* and *Indashyikirwa* (women) impacted all three physical IPV outcomes.

#### Physical and/or sexual IPV

Interventions differed in impact based on how outcomes were coded for the severity of physical and/or sexual IPV. *MAISHA CRT01* had no effect using the traditional any physical and/or sexual IPV measure (aOR:0.69, C.I.: 0.47-1.00). Our analysis finds that *MAISHA CRT01* reduced severe physical and/or sexual IPV when using the What Works measure (aOR:0.65, C.I.: 0.44-0.96). *Becoming One, Indashyikirwa* (women and men) and *Bandebereho,* on the other hand, showed an impact using both the What Works and severe physical and/or sexual IPV measures.

Male participants in the intervention group in the *SS-CF* study were less likely to report any perpetration of physical and/or sexual IPV (aRR:0.70, C.I.: 0.52-0.94) than the control group using the standard outcome measure. A reduction in severe physical and/or sexual IPV was found when using the What Works measure (aRR:0.70, C.I.: 0.52-0.94). However, there was no intervention impact when using the alternative severe physical and/or sexual IPV measure (aRR:0.73, C.I.: 0.54-1.01).

#### Emotional IPV

Several interventions differed in their measured impact on emotional IPV when comparing the standard approach to the two new indicators. In the *Becoming One* trial, participants in the intervention group showed a reduction in their experience of any emotional IPV compared to participants in the control group (aRR: 0.88, C.I.: 0.78-0.99). When assessing the impact of the intensity of emotional IPV, the intervention significantly reduced moderate and/or high-intensity emotional IPV (aRR: 0.74, C.I.: 0.57-0.96) but not high-intensity emotional IPV alone. On the other hand, the men’s UBL intervention had no effect on women’s reported experiences of emotional IPV as traditionally defined. However, the intervention did show an impact on both moderate and/or high-intensity emotional IPV (aOR: 0.73, C.I.: 0.56-0.96) and high-intensity emotional IPV only (aOR: 0.62, C.I.: 0.39-0.97). *MAISHA CRT01* had no impact on any emotional IPV, but the intervention did significantly decrease high-intensity emotional IPV only (aOR: 0.58, C.I.: 0.35-0.98). Bandebereho and Indashyikirwa (women) interventions demonstrated significant reductions in emotional IPV using all three outcomes.

There was no impact on men’s perpetration of emotional IPV using the traditional or new outcomes in *SS-CF* or *UBL*.

#### Difference in intervention impact by primary versus secondary prevention

Interventions differed in their impacts on primary vs secondary violence prevention when comparing the three outcomes of cessation, reduction and prevention. *Becoming One* significantly affected both women’s experiences of physical IPV and women’s experiences of sexual IPV using standard indicators. When intervention impacts were further assessed on primary vs secondary prevention outcomes, *Becoming One* was found to have prevented the new onset of physical IPV (aRR:1.01, C.I.: 1.01-1.10) among women who had not reported ongoing physical IPV at baseline. However, the intervention did not reduce ongoing physical violence or stop it completely. The opposite effects were seen for sexual IPV; intervention participants were more likely to report cessation of ongoing sexual IPV (aRR:1.17, C.I.: 1.02-1.34) at the endline than control group participants, but there was no impact on preventing new-onset sexual IPV. Similarly, *MAISHA CRT01* had an intervention effect on physical IPV using the standard indicators and prevented the new onset of physical IPV (aOR:1.80, C.I.: 1.13-2.87). The intervention was also effective in reducing ongoing physical IPV (aOR:2.54, C.I.: 1.05-6.19), but it did not impact cessation.

## Discussion

In this study, we reanalysed data from six trials to assess how different conceptualisations and coding of IPV variables can influence interpretations of the impact of an intervention. We compared standard outcome measures to new measures that reflect the severity of violence and whether interventions prevent new cases of IPV or reduce or stop ongoing violence. While we did not observe any clear trends *across* studies, we see important differences in intervention impact when comparing the standard outcome measures to the new ones. Importantly, in many trials, the traditional binary indicators masked some of the more subtle intervention effects, and the use of the new indicators allowed for a better understanding of the impacts of the interventions. At the same time, differences in results within studies between standard and new outcomes also differ across the six trials. While these results must be interpreted cautiously, given differences in intervention types, the underlying prevalence of violence, sociodemographic factors, sample sizes and other contextual differences across the trial sites, they can help us move toward a new approach to reporting multiple outcomes that allow us to dig deeper into the ‘impact’ of an intervention.

Several findings warrant further discussion. First, there was consistency in an intervention’s measured effectiveness across the standard and new outcome measures when the effect sizes were large. In two trials, *Bandebereho* and *Indashyikirwa* (women), the interventions showed consistent impacts on specific forms of IPV irrespective of whether the standard or new outcomes were assessed. For example, *Bandebereho* and *Indashyikirwa* (women) showed an effect when using the standard measure of any physical IPV and the new measures of moderate only physical and severe physical IPV. This was not the case for the other four trials. One possible explanation is that the effect size was greater in these two sites compared to other trials. In *Bandebereho*, for example, there was a 23 percentage point difference in reports of physical IPV between the intervention and control groups at the endline. The other trials, which showed differences in the magnitude of 0-4 percentage points, appeared to be more sensitive to how the outcomes were coded. For these four trials, assessing the differential impact on severe and moderate IPV captured effects that were not visible when using the standard dichotomous outcomes.

Our study raises several methodological issues. The first is identifying important differences in intervention impact based on the choice of outcome coding. In several trials, we found differences in the effectiveness of interventions by the severity of physical and emotional IPV. These differences can reflect *meaningful* differences in the impact of interventions on different severities of IPV or types of IPV (primary versus secondary) or methodological issues, including how we chose to code the variables or a lack of statistical power. It is challenging to ascertain a particular explanation. The rationale behind the differences in results has implications for the outcomes we use in IPV implementation research. Each of these concerns is discussed below in more detail.

We found differences in trial results by the intensity of IPV and type of prevention. Some of these results reflect meaningful differences in intervention impact. We found differences in the effects of *Becoming One* and *MAISHA CRT01* on primary and secondary prevention. These two interventions are very different. *Becoming One* uses faith leaders to work with couples, whereas *MAISHA CRT01* targeted women participating in microfinance programs. Differences in impact could be traced to the content of the program as well as the inclusion criteria or age groups for both interventions. Prior work in this area suggests that intervention strategies can have a differential impact by type of population. For example, interventions working with couples to identify and manage triggers of violence may be better suited to older, cohabiting couples who may be more invested in transforming their relationships as opposed to younger populations who may not yet be in long-term committed relationships and may be less invested in working with their partners to resolve relationship issues (Chatterji et al., 2020). Further research is needed to identify intervention strategies that may be more or less effective for primary or secondary violence prevention. To do this, studies need to specify a clear theory of change and pathways of impact for outcomes of interest.

Some of the differences in trial results can be attributed to underlying methodological decisions, including how variables were conceptualised and coded. For example, differences in results for the two measures of severe physical and/or sexual violence can be partially explained by differences in the impact on moderate-only physical IPV. The What Works measure of severe physical and/or sexual IPV includes moderate acts of physical IPV (slapping, pushing). In contrast, the other measure of severe physical and/or sexual IPV excludes moderate only acts of physical IPV. If trials had an impact on moderate only physical IPV but no impact on severe physical IPV (as seen in *MAISHA CRT01*), we see an impact using the What Works measure but not the other severe physical and/or sexual IPV measure. Conversely, the What Works measure fails to capture single acts of severe physical violence (because two acts are required) that occurred once (single acts are required to occur more than once in frequency to be captured), which could theoretically miss the impact of an intervention that affects severe violence by misclassifying single acts of severe violence to the reference group.

Similarly, our analyses on the differential impact of interventions on the intensity of emotional IPV highlight the methodological decision regarding the composition of the reference group. Interventions (*MAISHA CRT01, UBL* women) that did not impact any emotional IPV were found to affect moderate and/or high-intensity emotional IPV and/or high-intensity emotional IPV only. The reference group is the critical difference between the standard and the new outcomes. Compared to the traditional measure of any emotional IPV, individuals reporting low levels of emotional IPV are in the reference group of low or no emotional IPV, which potentially accounts for these differences in results between any emotional IPV and moderate and/or high-intensity emotional IPV or high-intensity emotional IPV only.

Nonetheless, these results on differences in impact by the severity of physical IPV and emotional IPV are relevant for the field of violence prevention as research has identified a dose-response relationship between the intensity of emotional IPV and adverse health outcomes (Follingstad & Rogers, 2013; Heise et al., 2019). Similarly, a few studies have highlighted poor health outcomes associated with more severe acts of physical IPV (Lacey & Mouzon, 2016; Signorelli et al., 2014). More cross-cultural research is needed to build on the work presented in this paper that tests the associations between IPV intensity and adverse health outcomes. Additionally, we need to develop a “gold standard” for measuring the severity of IPV. We chose to use WHO IPV items to establish measures of severity as these measures are widely used in IPV prevention research and the Demographic and Health Surveys conducted in over 60 countries to make it feasible for researchers to replicate our study. Researchers can also compare different strategies for measuring severity using cross-cultural data. It would be helpful to include data on injuries associated with different acts of violence to develop severity thresholds based on injury. Some acts categorised as moderate physical IPV, such as pushing, could theoretically result in serious injury in rare circumstances.

Our results on the differential impact of interventions on the severity of IPV can be used to refine the target population for interventions. Some interventions may be better suited to preventing more moderate forms of IPV rather than severe forms of IPV. More research is needed to unpack differences in these interventions, whether in content, strategy, delivery, or target populations. Lastly, we need to centre the voices of victims in this work on severity. There is a need for the co-production of qualitative research with victims to understand how they understand ‘severity’, what thresholds are meaningful for them, and whether these are based on the type of acts, frequency, the context in which the violence occurred, or the consequences. All acts of IPV are harmful to individuals, irrespective of severity. We undertook these analyses to see whether interventions differed in their impact by severity and our results show differences in effects by severity and intensity of IPV. These results underscore the importance of conducting these analyses while being mindful of the methodological issues involved.

These methodological issues have significant implications. When choosing outcomes, we need to ensure that our trials are adequately powered to detect differences based on the conceptualisation of the selected outcome. For example, the lack of adequate statistical power may have impacted some of our results. The analyses on primary and secondary prevention require us to stratify the sample into two subgroups based on baseline reporting of IPV; participants reporting ongoing IPV at baseline and participants reporting no IPV in a given period at baseline. Subgroup effects will be lower-powered than main effects due to each subgroup’s inherent smaller sample size. These analyses should be pre-specified to ensure an adequate sample size for each subgroup. We should also be cautious in over-interpreting these results, especially regarding null effects. Issues of statistical power are also relevant when choosing between binary and continuous outcome measures. We need to pay attention to the baseline prevalence of IPV when using a binary measure, as the variance increases with the baseline prevalence, reaching a maximum of 50%. The power to detect absolute and, to some extent, relative change varies with baseline prevalence. Programs that elect to use a binary measure should be conscious of the baseline prevalence in the setting and ensure they have adequate power to detect meaningful changes in violence. Lastly, when comparing traditional and new outcomes, such as moderate-only physical IPV and any physical IPV, it is essential to be cautious about overinterpreting differences based on statistical significance. There may be an overlap of confidence intervals, and a minor difference makes one variable ‘significant’ and the other not significant. For this reason, it is crucial to choose theoretically driven and contextually relevant outcomes.

We also found evidence of a gender difference in IPV reporting. In two trials, *Indashyikirwa* and *UBL*, we had data from couples. We found that a higher proportion of women reported experiencing severe physical IPV than men, who reported higher rates of perpetration of moderate-only physical IPV. Our results are similar to studies that have documented differences in rates of disclosure of IPV between men and women. We build on this literature by showing differences in disclosure rates by the severity of IPV. The motives behind differences in disclosure likely remain the same. A review of studies on gender discrepancy in IPV reporting found that factors affecting men’s underreporting of IPV perpetration in the U.S. and Spain included blaming their partner for provoking the violence to minimise their responsibility, fear of consequences and desire to avoid legal ramifications (Chan, 2011).

Overall, we have shown that the interpretation of an evaluation study can vary depending on the outcomes chosen and the way they are defined. Based on these observations, we offer the following recommendations for the violence prevention field. First, at the design stage, it is crucial to specify a theory of change and the hypothesised causal pathways by which a proposed intervention will achieve impact. Second, outcomes should be selected based on the theory of change, the specific aims of a given intervention, the underlying prevalence of violence, and the socio-cultural context of the field site. Where feasible, efforts should be made to include severity measures and determine impacts on primary and secondary violence prevention. Third, trialists should consider the methodological trade-offs associated with different outcome measures when choosing outcomes and designing the study. For example, the study should be designed to have a sufficient sample size to enable subgroup analyses on primary and secondary violence prevention. The choice of outcomes should be theoretically driven and trialists should pay careful attention to interpreting marginal differences between similar outcomes. Fourth, a baseline prevalence study should be conducted to refine outcomes based on preliminary data. This step will also make it possible to adjust for baseline variables that will increase the power of studies to detect outcome changes. Funding agencies must also prioritise the design stage for this data-driven decision-making. Fifth, we need more research to expand the knowledge base on the differential impact of interventions by the intensity of violence and type of prevention. We need to build on the results presented in this paper to unpack and identify different intervention strategies that can effectively target different intensities and types of violence. This type of analysis has not yet been carried out in the violence prevention field, and we must first develop a methodology to enable such research. This will allow us to refine our intervention strategies based on the baseline prevalence of different types and intensities of IPV. Lastly, funders and researchers must prioritise more IPV research on measurement issues. In this paper, we focused on outcome coding choices. More research is needed on other measurement issues, including statistical modelling, as there is no clear standard for best practices in modelling for binary or continuous variables in IPV implementation research.

### Limitations

This study has several limitations to be considered while interpreting the results. First, the prevalence rates presented in Table 1 are conservative as we compared endline rates as some sites did not collect baseline data. We did limit our analysis to prevalence in the control groups to avoid bias as far as possible. Second, given the cross-cultural nature of this study, it is difficult to draw conclusions about the differential impacts of interventions on different outcomes owing to differences in the type of intervention, the underlying prevalence of violence, sociodemographic factors, and sample sizes across sites. The exploratory analyses presented in this paper showcase the range of outcomes we can use in our evaluation studies to get us to think about the trade-offs of different approaches when choosing appropriate outcomes. Third, most of the novel outcomes assessed in this study were not pre-specified in the trial protocols. For example, although the *MAISHA CRT01* and *SS-CF* interventions impacted physical IPV, there was no effect on moderate-only physical IPV or severe physical IPV, which may be due to small sample sizes and lack of adequate power. This is also relevant for the subgroup analyses used to differentiate between the type of impact (primary vs secondary). Because these analyses were conducted post-hoc, trials may not be adequately powered for this kind of analysis, and these results should be considered exploratory.

## Conclusion

This study evaluated different approaches to coding outcome measures for assessing IPV prevention programs. Our results indicate that conclusions on whether a program is perceived “to work” are highly influenced by the IPV outcomes investigators choose to report and how they are measured and coded. Lack of attention to outcome choice and measurement could lead to prematurely abandoning strategies useful for violence reduction or missing essential insights into how programs may or may not affect IPV. As a young field, violence prevention must expand the range of outcomes tested to unpack differences between interventions, participants in the same intervention, and impact pathways for relevant subgroups.

## Data Availability

All data produced in the present study are available upon reasonable request to the authors. Please note that this manuscript presents secondary analysis of data from six trials.

**Appendix Table 3:**
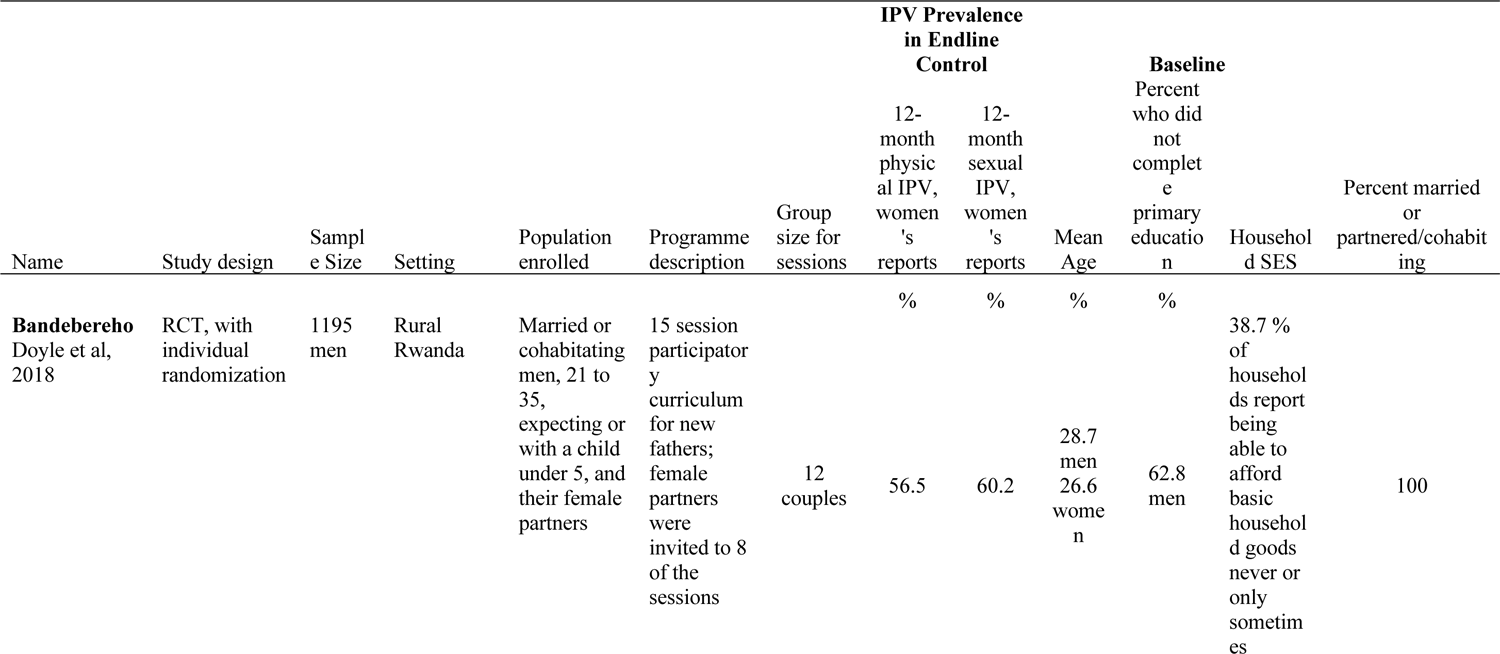
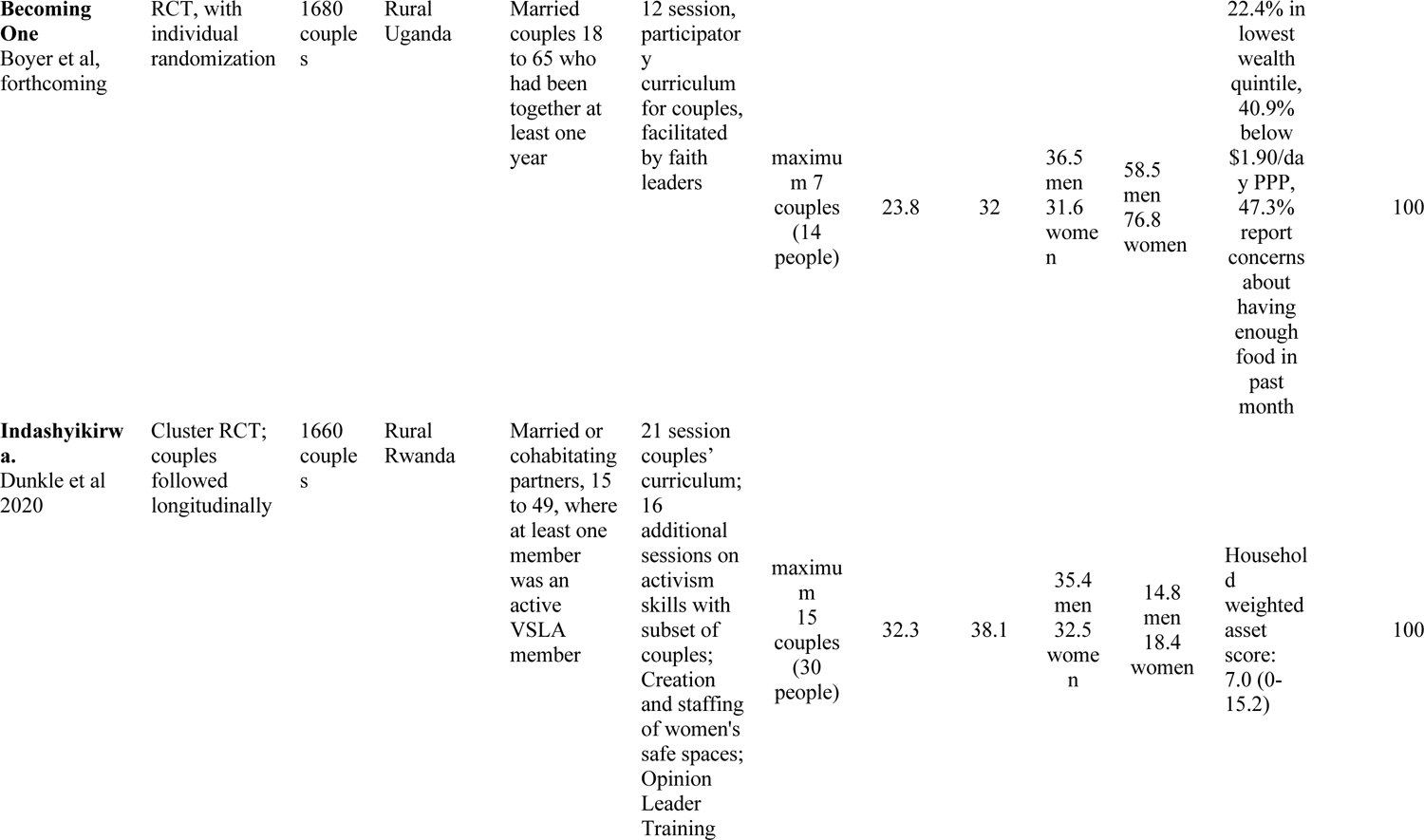
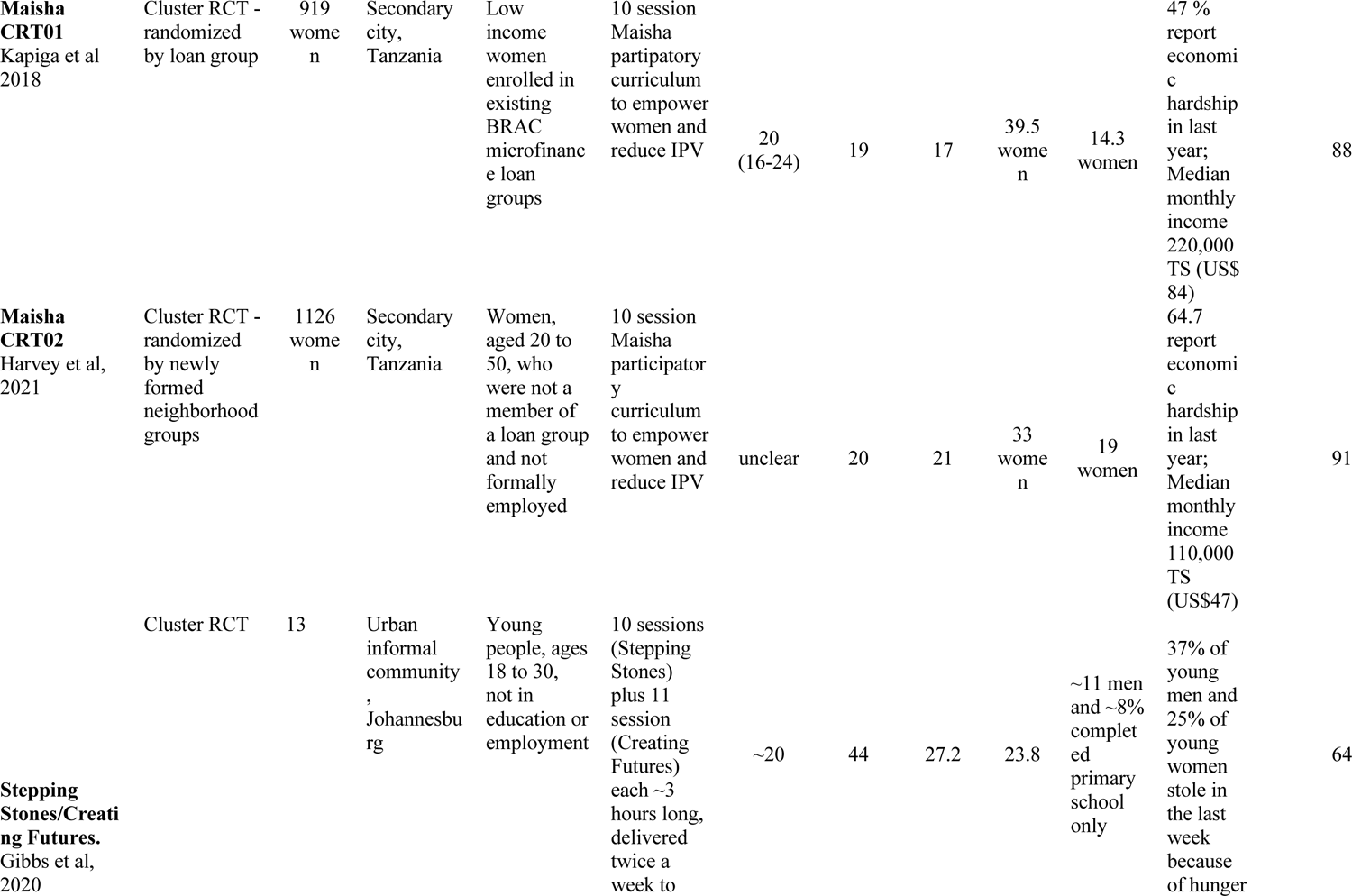
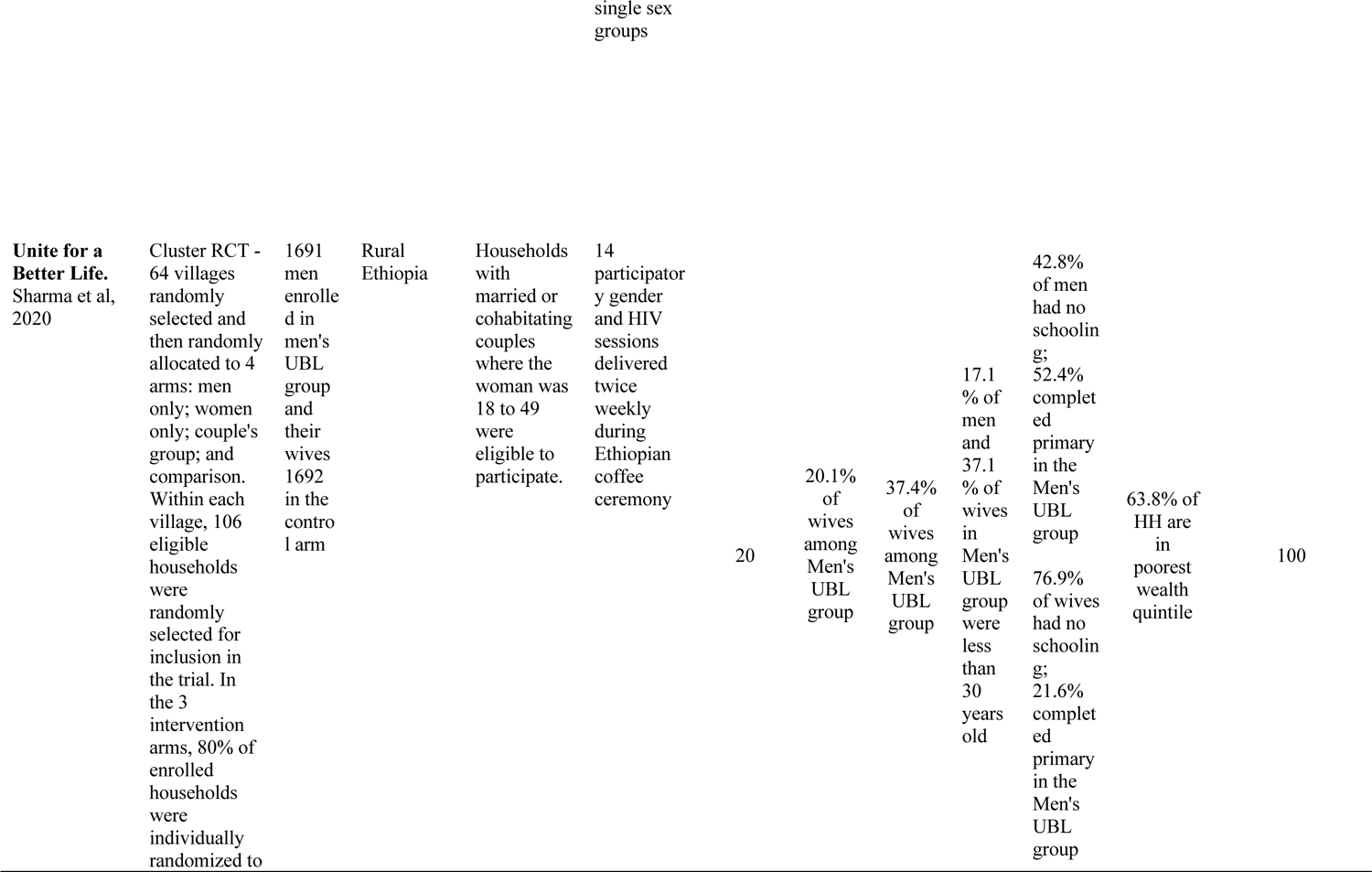
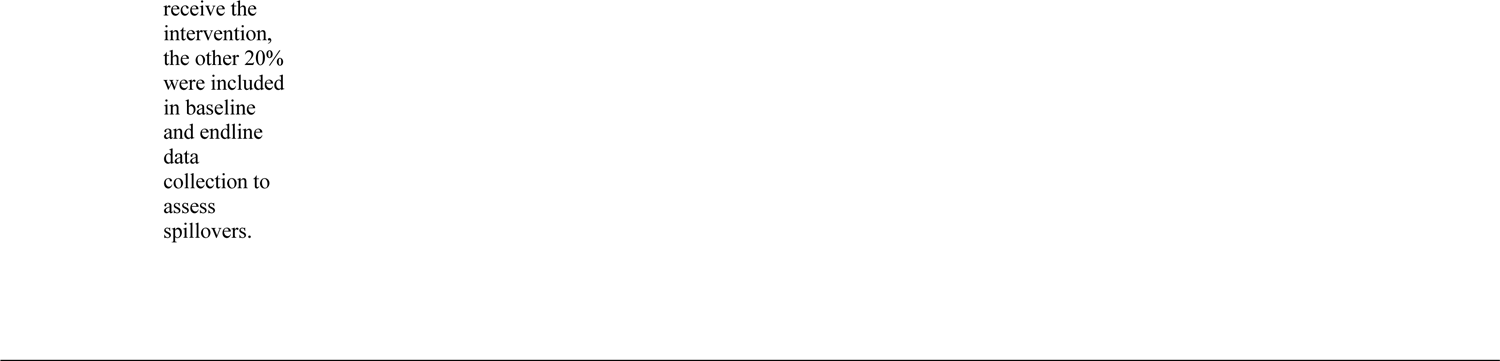
Description of Studies

**Appendix table 4:**
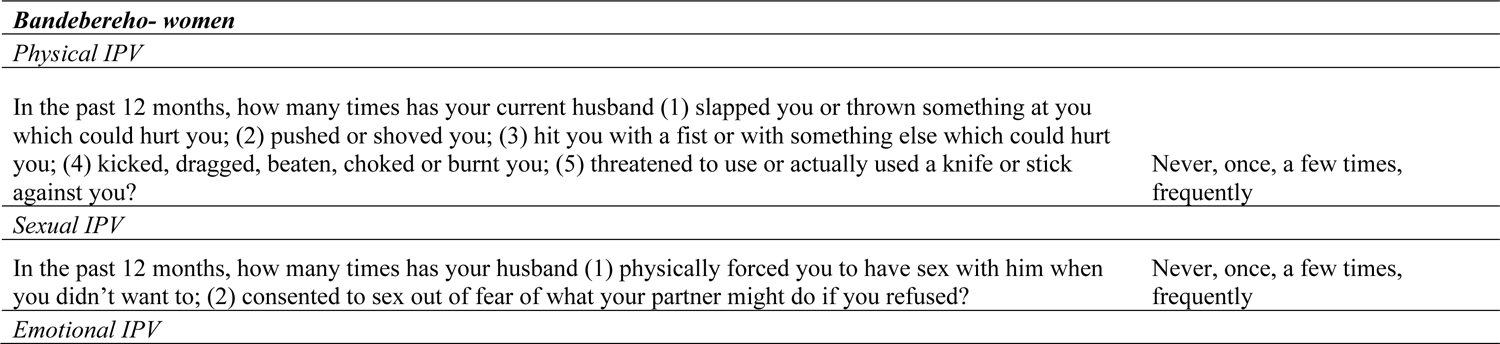
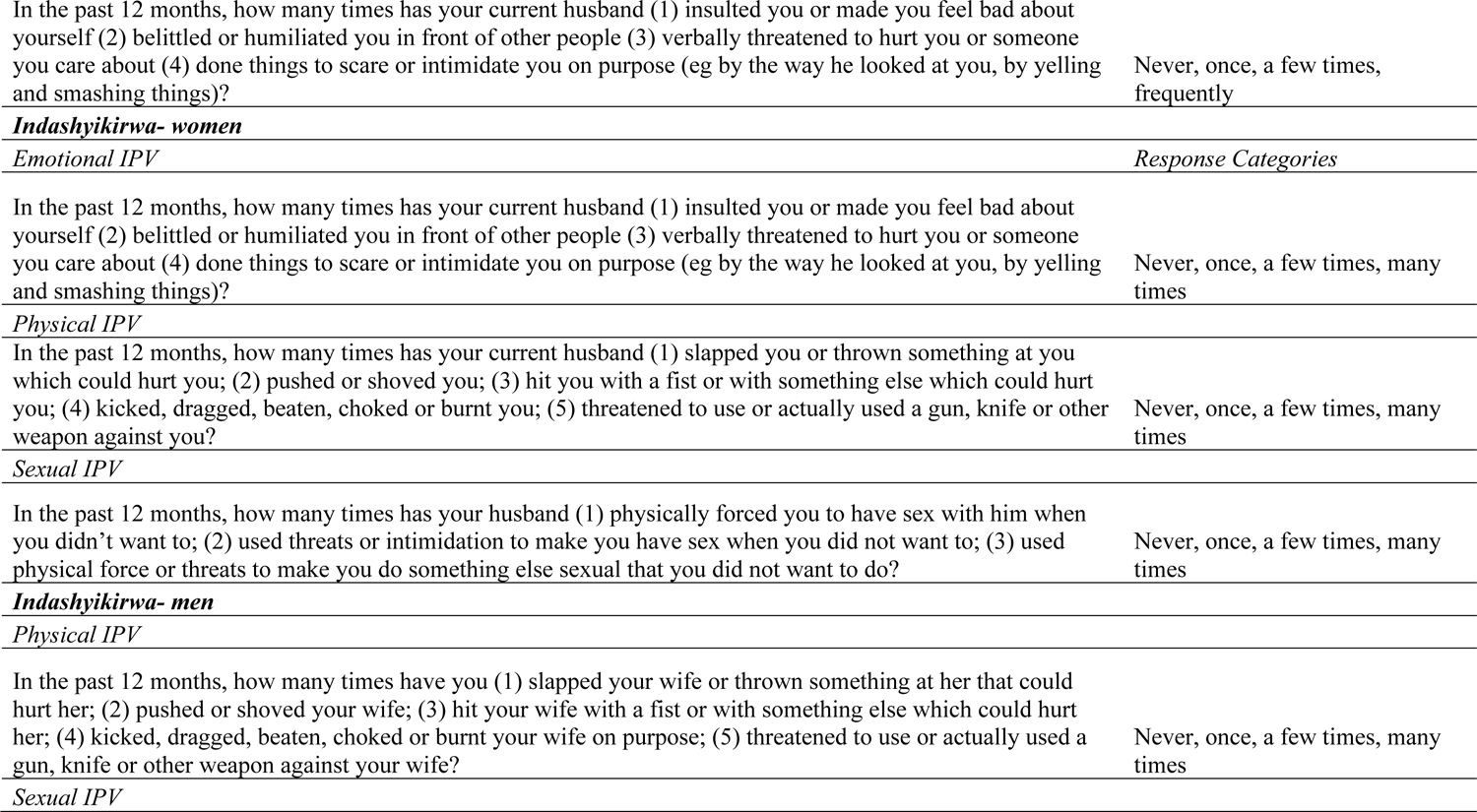
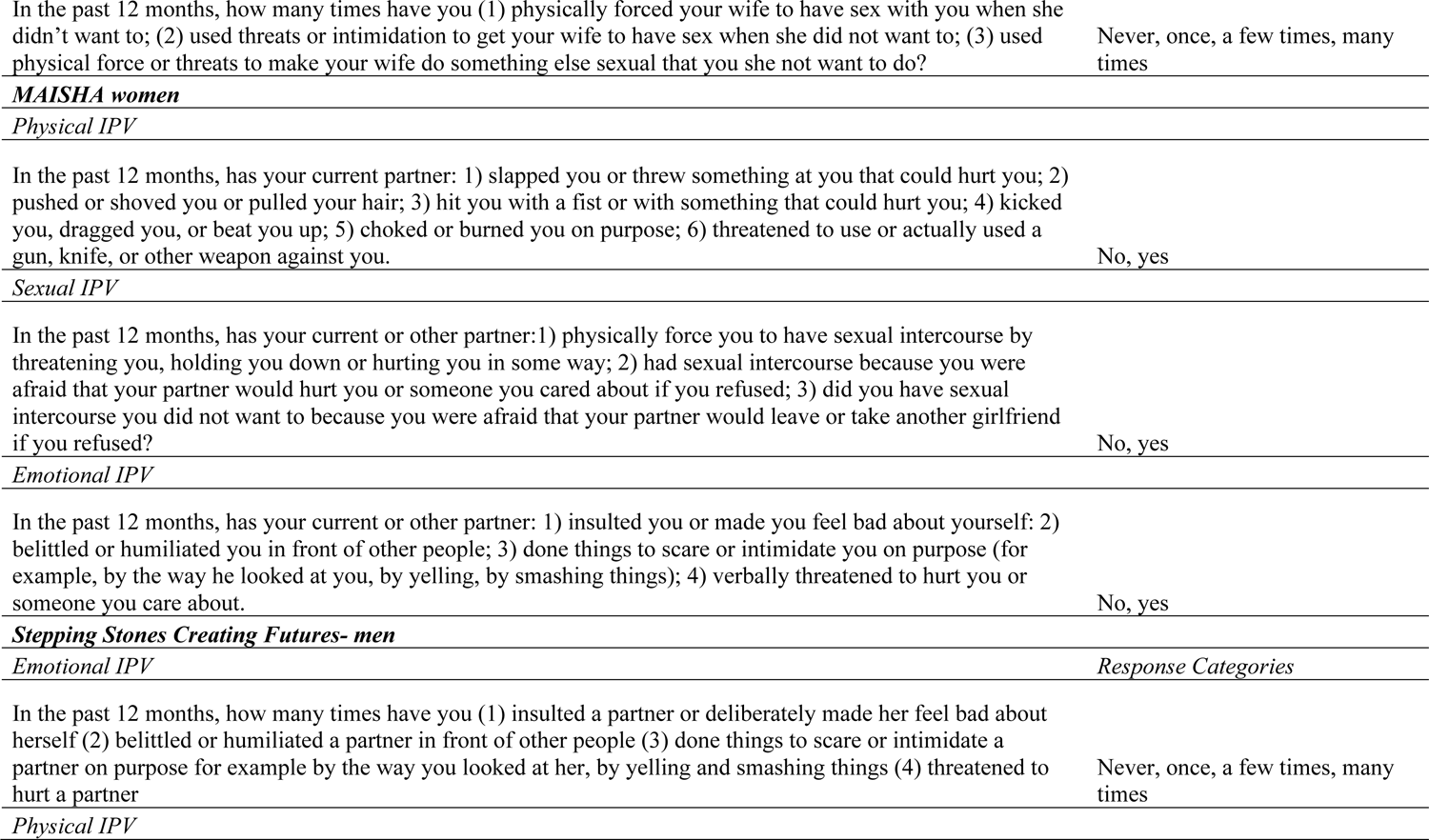
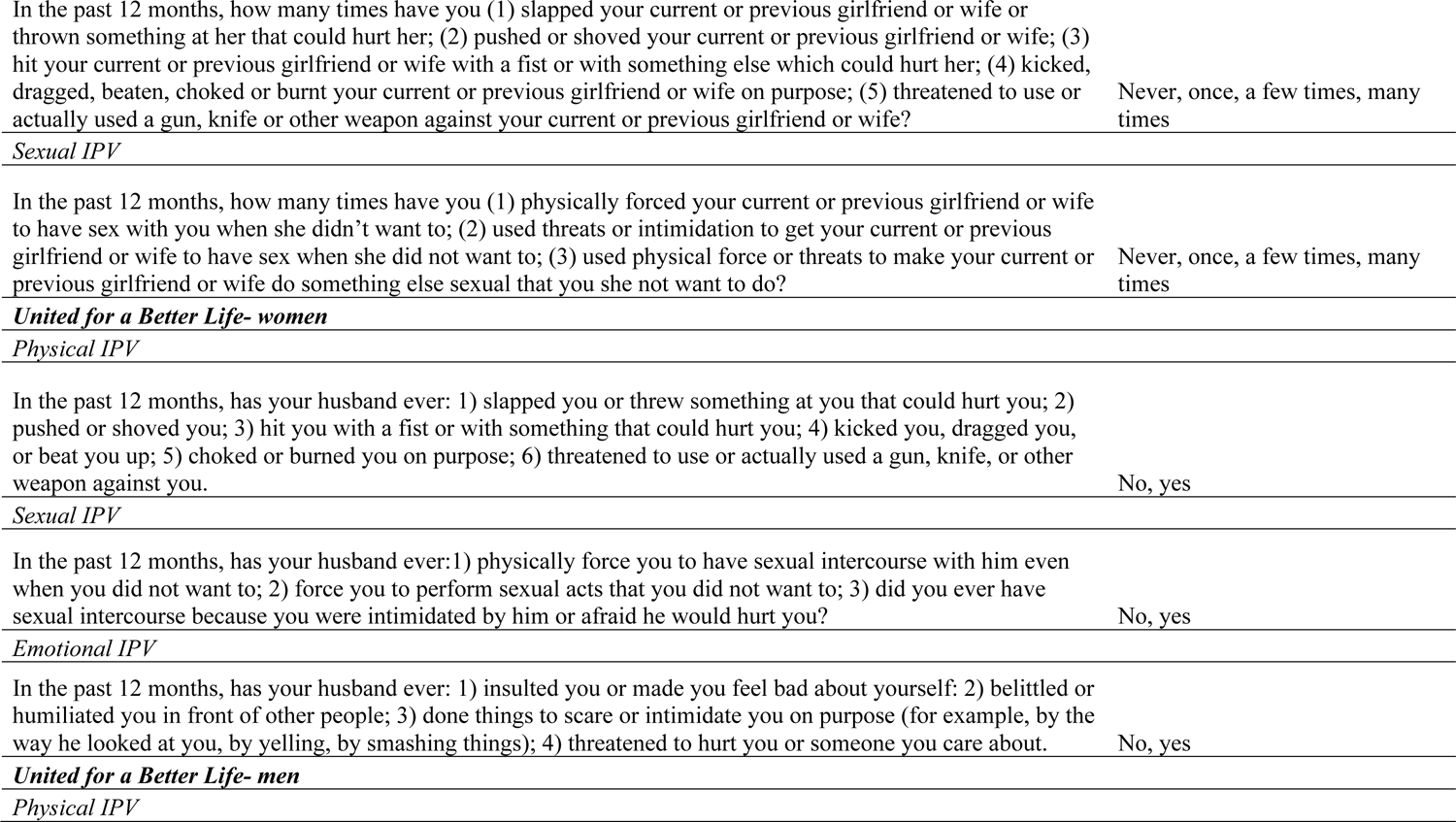
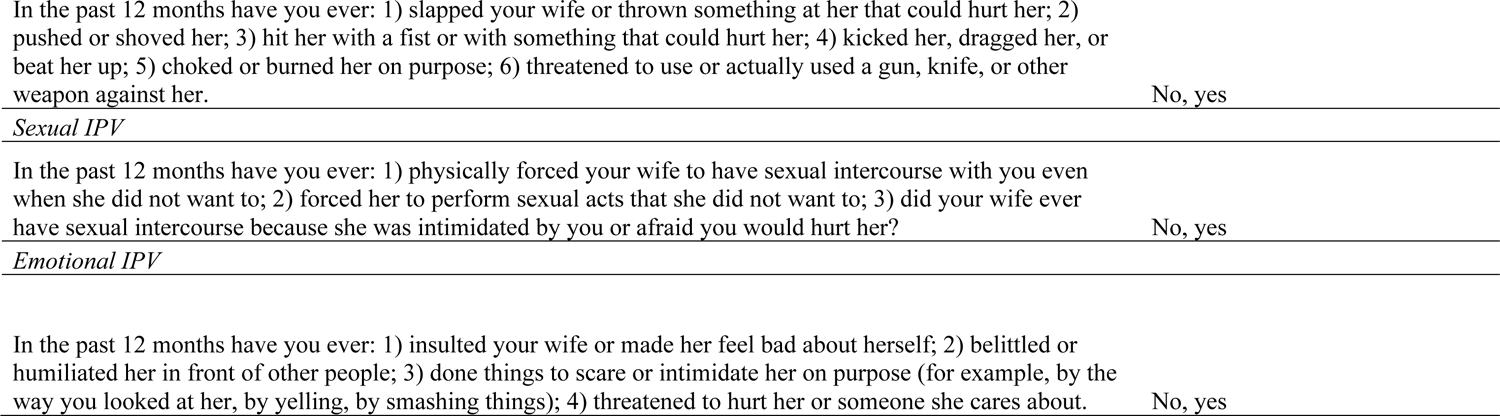
Items used to measure IPV across studies

## References

Abramsky, T., Devries, K. M., Michau, L., Nakuti, J., Musuya, T., Kyegombe, N., & Watts, C. (2016). The impact of SASA!, a community mobilisation intervention, on women’s experiences of intimate partner violence: secondary findings from a cluster randomised trial in Kampala, Uganda. J Epidemiol Community Health, 70(8), 818–825.

Bender, A. K. (2016, 2017/10/01). Ethics, Methods, and Measures in Intimate Partner Violence Research: The Current State of the Field. Violence Against Women, 23(11), 1382–1413. https://doi.org/10.1177/1077801216658977

Boyer, C., Levy Paluck, E., Annan, J., Nevatia, T., Cooper, J., Namubiru, J., Heise, L., & Lehrer, R. (2022, 2022/08/02). Religious leaders can motivate men to cede power and reduce intimate partner violence: Experimental evidence from Uganda. Proceedings of the National Academy of Sciences, 119(31), e2200262119. https://doi.org/10.1073/pnas.2200262119

Chan, K. L. (2011, 2011/03/01/). Gender differences in self-reports of intimate partner violence: A review. Aggression and Violent Behavior, 16(2), 167–175. https://doi.org/10.1016/j.avb.2011.02.008

Chatterji, S., Heise, L., Gibbs, A., & Dunkle, K. (2020). Exploring differential impacts of interventions to reduce and prevent intimate partner violence (IPV) on sub-groups of women and men: A case study using impact evaluations from Rwanda and South Africa. SSM - Population Health, 11, 100635. Retrieved 2020/08//, from https://doi.org/10.1016/j.ssmph.2020.100635

Clark, C. J., Bergenfeld, I., Cheong, Y. F., Kaslow, N. J., & Yount, K. M. (2022). Impact of measurement variability on study inference in partner violence prevention trials in low-and middle-income countries. Assessment, 10731911221095599.

Cunha, O. S., & Goncalves, R. A. (2018). Severe and Less Severe Intimate Partner Violence: From Characterization to Prediction. Violence and Victims(2), 235-250. https://doi.org/10.1891/0886-6708.VV-D-14-00033

Dickens, E., Augier, M., Sabet, S., Picon, M., & Rankin, K. (2019). Intimate partner violence prevention evidence gap map: 2018 update, 3ie Evidence Gap Map Report 8 (updated). https://doi.org/10.23846/EGM008

Doyle, K., Levtov, R. G., Barker, G., Bastian, G. G., Bingenheimer, J. B., Kazimbaya, S., Nzabonimpa, A., Pulerwitz, J., Sayinzoga, F., Sharma, V., & Shattuck, D. (2018). Gender-transformative Bandebereho couples’ intervention to promote male engagement in reproductive and maternal health and violence prevention in Rwanda: Findings from a randomized controlled trial. PLoS One, 13(4), e0192756–e0192756. https://doi.org/10.1371/journal.pone.0192756

Dunkle, K., Stern, E., Chatterji, S., & Heise, L. (2020). Effective prevention of intimate partner violence through couples training: a randomized controlled trial of <em>Indashyikirwa</em> in Rwanda. BMJ Global Health, 5(12), e002439. https://doi.org/10.1136/bmjgh-2020-002439

Follingstad, D. R. (2007). Rethinking current approaches to psychological abuse: Conceptual and methodological issues. Aggression and Violent Behavior, 12, 439–458.

Follingstad, D. R., & Rogers, M. J. (2013, 2013/08/01). Validity Concerns in Measuring Women’s and Men’s Intimate Partner Violence Report. Sex roles, 69(3), 149–167. https://doi.org/10.1007/s11199-013-0264-5

Gibbs, A., Washington, L., Abdelatif, N., Chirwa, E., Willan, S., Shai, N., Sikweyiya, Y., Mkhwanazi, S., Ntini, N., & Jewkes, R. (2020). Stepping Stones and Creating Futures intervention to prevent intimate partner violence among young people: cluster randomized controlled trial. Journal of Adolescent Health, 66(3), 323–335.

Grych, J., & Hamby, S. (2014). Advancing the measurement of violence: Challenges and opportunities. Psychology of violence, 4(4), 363.

Hamby, S. (2016). Self-report measures that do not produce gender parity in intimate partner violence: A multi-study investigation. Psychology of violence, 6(2), 323

Hamby, S. L. (2005). Measuring Gender Differences in Partner Violence: Implications from Research on Other Forms of Violent and Socially Undesirable Behavior. Sex roles, 52(11), 725–742. https://doi.org/10.1007/s11199-005-4195-7

Hardesty, J. L., Crossman, K. A., Haselschwerdt, M. L., Raffaelli, M., Ogolsky, B. G., & Johnson, M. P. (2015). Toward a standard approach to operationalizing coercive control and classifying violence types. Journal of Marriage and Family, 77(4), 833–843.

Harvey, S., Abramsky, T., Mshana, G., Hansen, C. H., Mtolela, G. J., Madaha, F., Hashim, R., Kapinga, I., Watts, C., & Lees, S. (2021). A cluster randomized controlled trial to evaluate the impact of a gender transformative intervention on intimate partner violence against women in newly formed neighbourhood groups in Tanzania. BMJ Global Health, 6(7), e004555.

Heise, L., Pallitto, C., García-Moreno, C., & Clark, C. J. (2019, 2019/12/01/). Measuring psychological abuse by intimate partners: Constructing a cross-cultural indicator for the Sustainable Development Goals. SSM - Population Health, 9, 100377. https://doi.org/10.1016/j.ssmph.2019.100377

Kapiga, S., Harvey, S., Mshana, G., Hansen, C. H., Mtolela, G. J., Madaha, F., Hashim, R., Kapinga, I., Mosha, N., & Abramsky, T. (2019). A social empowerment intervention to prevent intimate partner violence against women in a microfinance scheme in Tanzania: findings from the MAISHA cluster randomized controlled trial. The Lancet Global Health, 7(10), e1423–e1434.

Kerr-Wilson, A., Gibbs, A., E. M. F., Ramsoomar, L., Parke, A., Khuwaja, H., & Jewkes, R. (2019). A rigorous global evidence review of interventions to prevent violence against women and girls. https://www.whatworks.co.za/resources/evidence-reviews/item/693-a-rigorous-global-evidence-review-of-interventions-to-prevent-violence-against-women-and-girls

Keith, T., Hyslop, F., & Richmond, R. (2022). A Systematic Review of Interventions to Reduce Gender-Based Violence Among Women and Girls in Sub-Saharan Africa. Trauma, Violence, & Abuse, 15248380211068136. https://doi.org/10.1177/15248380211068136

Korman, L. M., Collins, J., Dutton, D., Dhayananthan, B., Littman-Sharp, N., & Skinner, W. (2008, 2008/03/01). Problem Gambling and Intimate Partner Violence. Journal of Gambling Studies, 24(1), 13–23. https://doi.org/10.1007/s10899-007-9077-1

Lacey, K. K., & Mouzon, D. M. (2016, 2016/09/01). Severe Physical Intimate Partner Violence and the Mental and Physical Health of U.S. Caribbean Black Women. Journal of Women’s Health, 25(9), 920–929. https://doi.org/10.1089/jwh.2015.5293

Lehrner, A., & Allen, N. E. (2014). Construct validity of the Conflict Tactics Scales: A mixed-method investigation of women’s intimate partner violence. Psychology of violence, 4(4), 477–490. https://doi.org/10.1037/a0037404

McNutt, L.-A., & Lee, R. (2000). Intimate Partner Violence Prevalence Estimation using Telephone Surveys: Understanding the Effect of Nonresponse Bias. American Journal of Epidemiology, 152(5), 438–441. https://doi.org/10.1093/aje/152.5.438

Ruiz-Pérez, I., Plazaola-Castaño, J., & Vives-Cases, C. (2007). Methodological issues in the study of violence against women. Journal of Epidemiology and Community Health, 61(Suppl 2), ii26. https://doi.org/10.1136/jech.2007.059907

Rowlands, I. J., Holder, C., Forder, P. M., Hegarty, K., Dobson, A. J., & Loxton, D. (2020, 2021/03/01). Consistency and Inconsistency of Young Women’s Reporting of Intimate Partner Violence in a Population-Based Study. Violence Against Women, 27(3-4), 359–377. https://doi.org/10.1177/1077801220908324

Sharma, V., Leight, J., Verani, F., Tewolde, S., & Deyessa, N. (2020). Effectiveness of a culturally appropriate intervention to prevent intimate partner violence and HIV transmission among men, women, and couples in rural Ethiopia: Findings from a cluster-randomized controlled trial. PLoS medicine, 17(8), e1003274. https://doi.org/10.1371/journal.pmed.1003274

Signorelli, M. S., Arcidiacono, E., Musumeci, G., Di Nuovo, S., & Aguglia, E. (2014, 2014/05/01). Detecting Domestic Violence: Italian Validation of Revised Conflict Tactics Scale (CTS-2). Journal of family violence, 29(4), 361-369. https://doi.org/10.1007/s10896-014-9594-5

Smith, C. A., Elwyn, L. J., Ireland, T. O., & Thornberry, T. P. (2010, 2010/03/01). Impact of Adolescent Exposure to Intimate Partner Violence on Substance Use in Early Adulthood. Journal of Studies on Alcohol and Drugs, 71(2), 219–230. https://doi.org/10.15288/jsad.2010.71.219

Straus, M. A. (1979). Measuring Intrafamily Conflict and Violence: The Conflict Tactics (CT) Scales. Journal of Marriage and Family, 41(1), 75–88. https://doi.org/10.2307/351733

Straus, M. A. (2017). The Conflict Tactics Scales and its critics: An evaluation and new data on validity and reliability. Routledge.

Straus, M. A., Hamby, S. L., Boney-McCoy, S. U. E., & Sugarman, D. B. (1996, 1996/05/01). The Revised Conflict Tactics Scales (CTS2): Development and Preliminary Psychometric Data. Journal of Family Issues, 17(3), 283-316. https://doi.org/10.1177/019251396017003001

Waltermaurer, E. (2005, 2005/04/01). Measuring Intimate Partner Violence (IPV): You May Only Get What You Ask For. Journal of interpersonal violence, 20(4), 501–506. https://doi.org/10.1177/0886260504267760

WalterMaurer, E. M., Ortega, C. A., & McNutt, L.-A. (2003, 2003/09/01). Issues in Estimating the Prevalence of Intimate Partner Violence: Assessing the Impact of Abuse Status on Participation Bias. Journal of interpersonal violence, 18(9), 959–974. https://doi.org/10.1177/0886260503255283

Yount, K. M., Bergenfeld, I., Mhamud, N., Clark, C. J., Kaslow, N. J., & Cheong, Y. F. (2022). Monitoring sustainable development goal 5.2: Cross-country cross-time invariance of measures for intimate partner violence. PLoS One, 17(6), e0267373. https://doi.org/10.1371/journal.pone.0267373

